# Life-Stage Heterogeneity in the Mental Health Treatment Gap: An Unsupervised Machine Learning Profiling of Symptomatic US Adults

**DOI:** 10.64898/2026.07.14.26358030

**Authors:** Wayne Lee Forday

## Abstract

**Background:** Despite a rising global psychiatric burden, a treatment gap persists where the majority of symptomatic individuals remain unmedicated. Traditional epidemiological analyses treat this untreated population as a single, uniform block, obscuring specific barriers to care. This study uses an unsupervised machine learning pipeline to identify distinct socio-behavioural and biological sub-populations within the untreated cohort to guide targeted public health interventions.

**Methods:** Data were pooled from the 2015-2018 National Health and Nutrition Examination Survey (NHANES) cycles (*N* = 11,848 total adult respondents). A symptomatic cohort of 3,075 individuals experiencing daily or weekly anxiety or depression symptoms was isolated, excluding severe liver pathology outliers (GGT ≥ 80 U/L). A 22-feature matrix combining continuous clinical biomarkers (systolic blood pressure, waist circumference, HbA1c) and categorical social variables was projected using Factor Analysis of Mixed Data (FAMD). Latent sub-populations were identified via Gaussian Mixture Modelling (GMM), optimized by the Bayesian Information Criterion (BIC).

**Results:** The broad baseline population revealed a substantial mental health burden, with 30.4% reporting active psychiatric symptoms, of whom 71.6% were entirely unmedicated. The GMM pipeline successfully isolated three distinct sub-populations (*k* = 3) separated by age, clinical strain, and treatment rates:

- **Cluster 0 (Mature Adults, mean age 55.03):** high psychiatric severity (34.1% severe untreated), central obesity, and hypertensive strain (135.82 mmHg), with 64.1% untreated despite frequent primary care contact;
- **Cluster 1 (Working Professionals, mean age 38.38):** highly educated, female-dominated (70.5%), with 77.7% untreated driven by moderate distress;
- **Cluster 2 (Emerging Youth, mean age 18.49):** a highly vulnerable late-adolescent group with a staggering 90.2% untreated rate.

**Conclusion:** The unmedicated symptomatic population is highly diverse and segmented by life stage. These profiles show that the treatment gap is driven by age-specific barriers, specifically workforce-age symptom masking and late-adolescent developmental transitions. Closing this deficit requires shifting from uniform public health approaches toward targeted interventions, such as digital peer support networks for youth and integrated primary care screenings for older adults.

## Introduction

Mental health disorders represent one of the most pervasive public health challenges of the twenty-first century. Symptoms of anxiety and depressive disorders contribute substantially to the global burden of disease, driving widespread functional impairment, diminished quality of life, and significant socioeconomic strain. Yet, despite the availability of validated clinical interventions, a well-documented paradox defines modern psychiatric epidemiology: the majority of individuals suffering from active psychological distress remain disconnected from formal treatment networks. This discrepancy—frequently termed the mental health “treatment gap”—presents a substantial barrier to public health systems worldwide.

Historically, epidemiological frameworks have analyzed this untreated population as a single, homogeneous block. Public health strategies have consequently relied on uniform, population-wide campaigns designed to lower costs or expand general clinical access. While well-intentioned, these one-size-fits-all approaches may fail because they ignore the complex, multi-dimensional barriers that prevent different groups from seeking care. The reasons an individual remains untreated are closely tied to their stage of life, intersecting with socioeconomic constraints, coping strategies, and physical health.

To bridge this gap, public health research may benefit from shifting from broad population averages toward more granular, data-driven subclass targeting. This study addresses this need by applying an unsupervised machine learning pipeline to a large, nationally representative cohort of symptomatic adults. By examining the interactions between clinical biomarkers and social infrastructure variables, this paper aims to identify the sub-populations driving the unmedicated mental health burden, providing a framework for targeted, life-stage-specific community interventions.

## Literature Review

### Macro Scale of the Unmet Treatment Gap

The scope of the global mental health treatment gap is extensively documented across diverse clinical and cultural landscapes. Large-scale epidemiological surveys consistently show that structural, financial, and attitudinal barriers prevent the majority of symptomatic individuals from accessing psychiatric care. This structural deficit is not unique to specific regions or healthcare models; it represents a cross-cultural public health reality. For example, recent community-based research in Asian urban centers, such as the study by Teo et al. (2026) published in the *Singapore Medical Journal*, reported that 77% of adults living with active symptoms of anxiety or depression had abstained from seeking formal care.

While macro-level surveys are highly valuable for establishing baseline prevalence metrics, they are inherently limited by their reliance on broad population averages. When data are aggregated into a single cohort, the unique barriers experienced by distinct demographic groups may become obscured. In addition, traditional public health studies often rely on small sample sizes—the Teo et al. framework, for instance, was limited to 350 participants—which may limit the identification of more nuanced sub-populations.

### Somatic-Psychiatric Gradient and Clinical Disconnection

A limitation in modern mental health literature is the historical separation between psychiatric symptoms and objective physical pathology. Many large-scale mental health studies rely primarily on self-reported questionnaire data, omitting physical examinations and objective biochemical laboratory metrics. This data gap overlooks an important biological mechanism: chronic psychiatric distress is associated with prolonged hypothalamic-pituitary-adrenal (HPA) axis activation, which may contribute to systemic physical changes across the lifespan. Chronic stress induces HPA axis dysregulation, autonomic nervous system imbalance, and abnormal inflammatory responses, which together drive the development of multiple psychosomatic diseases.

Untreated mental illness is associated with somatic health outcomes, including visceral obesity, cardiovascular strain, glycemic decline, and hepatic enzyme elevation. The concept of allostatic load—multi-system physiological dysregulation—captures how psychiatric disorders, such as depression, are associated with dysregulation across immune, HPA-axis, and autonomic nervous systems. Because mature adults experiencing these somatic symptoms interact frequently with primary care networks for chronic disease monitoring, their continued non-treatment represents a missed clinical opportunity. Primary care clinicians may treat physical biomarkers in isolation, potentially missing underlying psychiatric distress. To fully understand the treatment gap, analytical models may benefit from integrating objective biochemical laboratory data alongside social indicators.

### Workforce Masking and Adolescent Transition Barriers

The barriers preventing individuals from accessing care vary substantially across different life stages. In working-age cohorts, the treatment gap is often associated with “behavioural masking”—the phenomenon where individuals manage persistent symptoms while maintaining external functioning. High-functioning professionals may manage persistent, moderate psychiatric symptoms in clinical isolation, maintaining careers, families, and metabolic baselines while avoiding formal medical networks due to time constraints and concerns about professional stigma. The Singaporean study by Teo et al. (2026) found that white-collar managerial professionals expressed greater interest in informal peer support than non-managerial workers, suggesting that workplace hierarchy and professional stigma influence help-seeking preferences.

Conversely, individuals transitioning into early adulthood face different structural obstacles. Late adolescents frequently age out of paediatric or school-based support networks, entering a critical developmental phase where they may lack independent economic resources, stable social structures, and the healthcare navigation experience required to access adult medical systems. Young adults consistently report lower formal service utilization despite high symptom burden, and mental health problems in college-aged populations are associated with significant academic and functional impairment. Broad population models may flatten these distinct age-specific patterns, combining working professionals and transitioning youth into a singular average.

### Methodological Advancements via Unsupervised Machine Learning

To overcome the limitations of standard linear statistics, recent public health research has increasingly turned to unsupervised machine learning. Traditional partitioning algorithms, such as Principal Component Analysis (PCA) and k-means clustering, are structurally unsuited for complex epidemiological datasets because they cannot handle mixed data types and may force populations into artificial, equal-sized spherical boundaries.

By applying Factor Analysis of Mixed Data (FAMD), researchers can balance the geometric weight of continuous clinical markers and categorical social variables simultaneously, preventing either data type from dominating the latent space. FAMD is specifically designed for mixed datasets, weighting categorical and continuous variables to balance their contributions to the principal components. When paired with a probabilistic Gaussian Mixture Model (GMM) utilizing a full covariance matrix, the algorithm accommodates flexible, overlapping, and elliptical population densities.

This approach is particularly well-suited for preserving small, high-density sub-populations—such as adolescent transition bands—that may be obscured by less flexible methods.

## Methods

### Study Population and Data Source

This study used data from the National Health and Nutrition Examination Survey (NHANES), a nationally representative cross-sectional survey conducted by the National Center for Health Statistics (NCHS) of the Centers for Disease Control and Prevention (CDC). NHANES employs a complex, multistage probability sampling design to select participants representative of the civilian non-institutionalized U.S. population.

Data were pooled from two consecutive NHANES cycles spanning 2015-2018 (survey years 2015-2016 and 2017-2018). As this was a secondary analysis of publicly available, fully de-identified data, institutional review board approval was not required. All data were obtained directly from the public CDC NHANES repository.

### Cohort Selection and Case Definitions

The final analytical sample was established using a three-tier screening framework:

1. **Broad Population Baseline:** A total of 11,848 adult respondents aged 18 years and older with valid mental health questionnaire data were retained to establish nationwide baseline prevalence metrics.
2. **Symptomatic Filter:** Clinical cases were defined based on the frequency of self-reported anxiety or depression symptoms. Participants reporting symptoms “Daily” or “Weekly” were classified as symptomatic. This sub-cohort comprised 3,607 individuals (30.4% of the total sample).
3. **Final Analytical Cohort:** To prevent severe pathological liver conditions from distorting latent variance metrics during dimensionality reduction, individuals with gamma-glutamyl transferase (GGT) levels ≥ 80 U/L were excluded. This quality control step yielded a final analytical cohort of *N* = 3,075 symptomatic individuals (26.0% of the original sample population).

### Primary Outcome and Validation Check

The primary outcome was untreated status, defined as meeting the symptomatic cohort criteria while simultaneously answering “No” to current use of prescription medication for anxiety (NHANES variable DLQ110) and depression (NHANES variable DLQ150). Participants who reported active compliance with either medication regimen were classified as treated.

To validate the self-reported symptom tracking framework, a concordance check was performed against the standardized 9-item Patient Health Questionnaire (PHQ-9).

The self-reported depression indicator (NHANES variable DLQ150) demonstrated high diagnostic agreement, achieving an 89.3% concordance rate with a PHQ-9 threshold score of ≥ 10 (representing moderate-to-severe depression).

### Data Extraction and Harmonization

Data were extracted from multiple NHANES component files using the pandas library (Python 3.8). Features were categorized as continuous or categorical to support a comprehensive biopsychosocial analytical framework.

#### Sociodemographic and Infrastructure Features

Demographic component files (DEMO) provided data on core socioeconomic and structural variables.

##### Continuous Variables

- Age (years)
- Income_PIR: Poverty Income Ratio, a continuous measure of household income relative to the federal poverty line
- Household_Size: Number of persons in the household

##### Categorical Variables

- Gender: Male/Female
- Race_Ethnicity: Classified as White, Hispanic, Black, or Other based on the RIDRETH1 variable
- Education: Categorized as Less than High School, High School Graduate, Some College/Higher, or Not Administered/Age < 20
- Marital_Status: Categorized as Married/Partnered, Never Married, Divorced/Separated, Widowed, or Not Administered/Age < 20
- AlcoholCategory: Categorized as Non-drinker, Occasional Drinker, Moderate Drinker, Heavy Drinker, Binge/High Intensity Drinker, or Not Administered/Age < 20

For individuals under 20 years of age, NHANES administrative branching logic automatically skips the Education and Marital_Status questionnaires. Rather than dropping these records or using statistical imputation, missing values were preserved as an explicit descriptive category: “Not Administered / Age < 20”.

### Behavioural and Lifestyle Features

Behavioural and lifestyle variables were derived from dietary, smoking, and physical activity questionnaires.

#### Continuous Variables

- Total_Calories: Derived from the Day 1 Total Nutrient Intakes file (DR1TOT) via a 24-hour dietary recall interview
- Sleep_Hours: Self-reported daily sleep duration
- Sedentary_Activity: Self-reported daily sitting time, converted to hours per day (derived from NHANES physical activity questionnaire variables)
- Activity_Days: Sum of days per week a participant reported engaging in moderate or vigorous physical activity (derived from NHANES PAQ variables)

#### Categorical Variables

- Smoking_Status: Classified as Never Smoker, Former Smoker, Current Light Smoker (≤ 10 cigarettes/day), Current Moderate Smoker (11-20 cigarettes/day), Current Heavy Smoker (> 20 cigarettes/day), or Not Administered/Age < 20

### Anthropometric and Cardiovascular Features

Physical examination measures were extracted from NHANES Examination components.

#### Continuous Variables

- Waist_Circumference (cm): Measured at the iliac crest using standardized protocols by trained health technicians
- sbp_mean (mmHg): Mean systolic blood pressure derived from up to three consecutive readings obtained using a mercury sphygmomanometer. To adjust for medication-induced masking, 10 mmHg was added to the measured SBP for participants reporting current use of antihypertensive medication. This correction is a standard approach to estimate pre-treatment blood pressure levels, as the average reduction in systolic blood pressure achieved by most antihypertensive pharmacotherapy is approximately 10 mmHg.

### Biochemical and Metabolic Features

Metabolic and organ-function markers were extracted from NHANES Laboratory components.

#### Continuous Variables

- HbA1c (%): Glycemic status from the Glycohemoglobin file (GHB)
- GGT_Liver_Enzyme (U/L): Gamma-glutamyl transferase from the Biochemistry Profile file (BIOPRO)
- Alkaline_Phosphatase (U/L): Alkaline phosphatase from BIOPRO
- Albumin (g/L): Serum albumin from BIOPRO
- Uric_Acid (*μ*mol/L): Blood uric acid from BIOPRO
- Dietary cholesterol (mg): Total daily dietary cholesterol intake from the First Day Dietary Interview file (DR1TOT)

#### Skeletal Muscle Index

Skeletal muscle mass (SM) was estimated using sex-specific anthropometric equations validated by Kim et al., incorporating weight (W, kg), height (H, cm), age (A, years), waist circumference (WC, cm), and race (R):

- **Males:** SM = 0.47(W) + 0.03(H) + 0.012(A) − 0.001(A^2^) − 0.29(WC) + 1.6(R) + 13.5
- **Females:** SM = 0.25(W) + 0.09(H) − 0.11(A) + 0.0005(A^2^) − 0.06(WC) + 2.0(R) − 4.5

Race was coded as 0 for White and 1 for Black, consistent with the original validation sample. Hispanic participants were conservatively coded as White (*R* = 0), as the equations were not validated for this population. Participants of “Other” race were excluded from muscle mass estimation. A physiological plausibility filter excluded estimates outside the 0-100 kg range. The final feature was expressed as the skeletal muscle index (SMI), calculated as SM divided by height squared (kg/m^2^), consistent with appendicular skeletal muscle index conventions.

### Data Quality Control and Advanced Preprocessing

#### Outlier Filtration and Truncation

All data underwent quality control procedures in which NHANES-specific missing codes (e.g., 7, 9, 99, 999) were systematically recoded to true missing values (NaN). Exploratory extreme-value analysis using the interquartile range (IQR) method identified substantial right-skewing in biochemical features, particularly for GGT_Liver_Enzyme, where severe clinical outliers extended up to 1,681.0 U/L. Because these extreme pathological outliers can heavily skew latent variance projections during dimensionality reduction, a truncation threshold was applied, restricting the final modelling cohort to individuals with GGT levels < 80 U/L. No other continuous variables required truncation based on the IQR criterion.

#### Dimensionality Reduction and Unsupervised Profiling

To prepare the final feature matrix for latent class analysis, a structured data preprocessing pipeline was implemented. Missing values in categorical features were explicitly coded as “Unknown” to preserve structural completeness and cast to categorical data types. Continuous missing values were imputed using the median (scikit-learn SimpleImputer with strategy=‘median’). The complete-case feature matrix—comprising 16 continuous and 6 categorical variables—was then advanced to dimensionality reduction and probabilistic cluster modelling.

**Table 1.** Variable completeness prior to imputation in the symptomatic sample (*N* = 3, 075)

| **Variable** | **Non-Missing (n)** | **% Complete** | **Pipeline Type** |
| --- | --- | --- | --- |
| Age | 3,075 | 100.0% | Continuous |
| Gender | 3,075 | 100.0% | Categorical |
| Race_Ethnicity | 3,075 | 100.0% | Categorical |
| GGT_Liver_Enzyme | 3,075 | 100.0% | Continuous |
| Albumin | 3,075 | 100.0% | Continuous |
| Household Size | 3,075 | 100.0% | Continuous |
| Alkaline Phosphatase | 3,074 | 99.9% | Continuous |
| Uric Acid | 3,073 | 99.9% | Continuous |
| Smoking_Status | 3,064 | 99.6% | Categorical |
| HbA1c | 3,056 | 99.4% | Continuous |
| Sedentary_Activity | 3,033 | 98.6% | Continuous |
| sbp_mean | 2,972 | 96.6% | Continuous |
| Activity_Days | 2,934 | 95.4% | Continuous |
| Waist Circumference | 2,906 | 94.5% | Continuous |
| Marital_Status | 2,890 | 94.0% | Categorical |
| Education | 2,889 | 93.9% | Categorical |
| Total Calories | 2,849 | 92.7% | Continuous |
| Dietary Cholesterol | 2,849 | 92.7% | Continuous |
| Income_PIR | 2,748 | 89.4% | Continuous |
| Muscle_Mass_Height_Ratio | 2,525 | 82.1% | Continuous |
| AlcoholCategory | 2,324 | 75.6% | Categorical |
| Sleep_Hours | 2,253 | 73.3% | Continuous |

## Statistical Modelling and Unsupervised Cluster Analysis

### Dimensionality Reduction via Factor Analysis of Mixed Data (FAMD)

Because the final analytical feature matrix comprised a complex mixture of 16 continuous variables and 6 categorical variables, traditional dimensionality reduction techniques—such as standard Principal Component Analysis (PCA) for continuous data or Multiple Correspondence Analysis (MCA) for categorical data—were mathematically inappropriate. To preserve the variance of both data types simultaneously, we applied Factor Analysis of Mixed Data (FAMD). FAMD is specifically designed for mixed datasets, weighting categorical and continuous variables to balance their contributions to the principal components.

FAMD normalizes continuous variables to a mean of 0 and unit variance, while projecting categorical indicators using a scaled disjunction matrix. This mathematical integration balances the geometric weight of clinical biomarkers and social infrastructure variables, ensuring neither data type dominates the latent space due to measurement scale differences. To reduce background noise while capturing the core variance of the 22 features, the top five orthogonal dimensions (*n* = 5 principal components) were extracted and retained for downstream cluster modelling.

### Latent Class Identification via Gaussian Mixture Modelling (GMM)

To discover natural typologies within the symptomatic cohort, a Gaussian Mixture Model (GMM) was applied to the five-dimensional FAMD coordinate space. Unlike rigid, distance-based partitioning algorithms (such as k-means) that force populations into artificial, equal-sized spherical clusters, GMM treats clustering as a density estimation problem. By optimizing a full covariance matrix, the GMM allows for flexible, overlapping, oblong, and elliptical cluster boundaries of varying densities and volumes. This mathematical flexibility accommodates the soft, probabilistic boundaries inherent to human life-stage transitions and safely isolates smaller, high-density subgroups that traditional sphere-fitting models would absorb or obscure.

### Model Selection and Cluster Optimization

The optimal number of latent clusters was determined through a hyperparameter grid search across a component range of *k* = 2 to *k* = 6. Model fit was evaluated using the Bayesian Information Criterion (BIC) and the Akaike Information Criterion (AIC) to balance model likelihood against parameter complexity.

The optimal cluster solution was established at *k* = 3 components. This selection was dictated by a distinct “elbow” inflection point on the BIC curve, beyond which adding further components yielded diminishing returns in log-likelihood alongside an increased risk of overparameterization. To evaluate the spatial separation and structural integrity of the optimized 3-cluster framework, final GMM assignments were projected and visualized in a two-dimensional scatter plot using the coordinates of the first two principal components of the FAMD transformation.

## Results

### Baseline Characteristics of the Symptomatic Cohort

The final analytical cohort consisted of *N* = 3,075 symptomatic individuals derived from the 2015-2018 NHANES cycles. The population demonstrated a substantial psychiatric burden that intersected with notable socioeconomic constraints, lifestyle risk factors, and underlying cardiometabolic strain (Table 2).

**Table 2.**
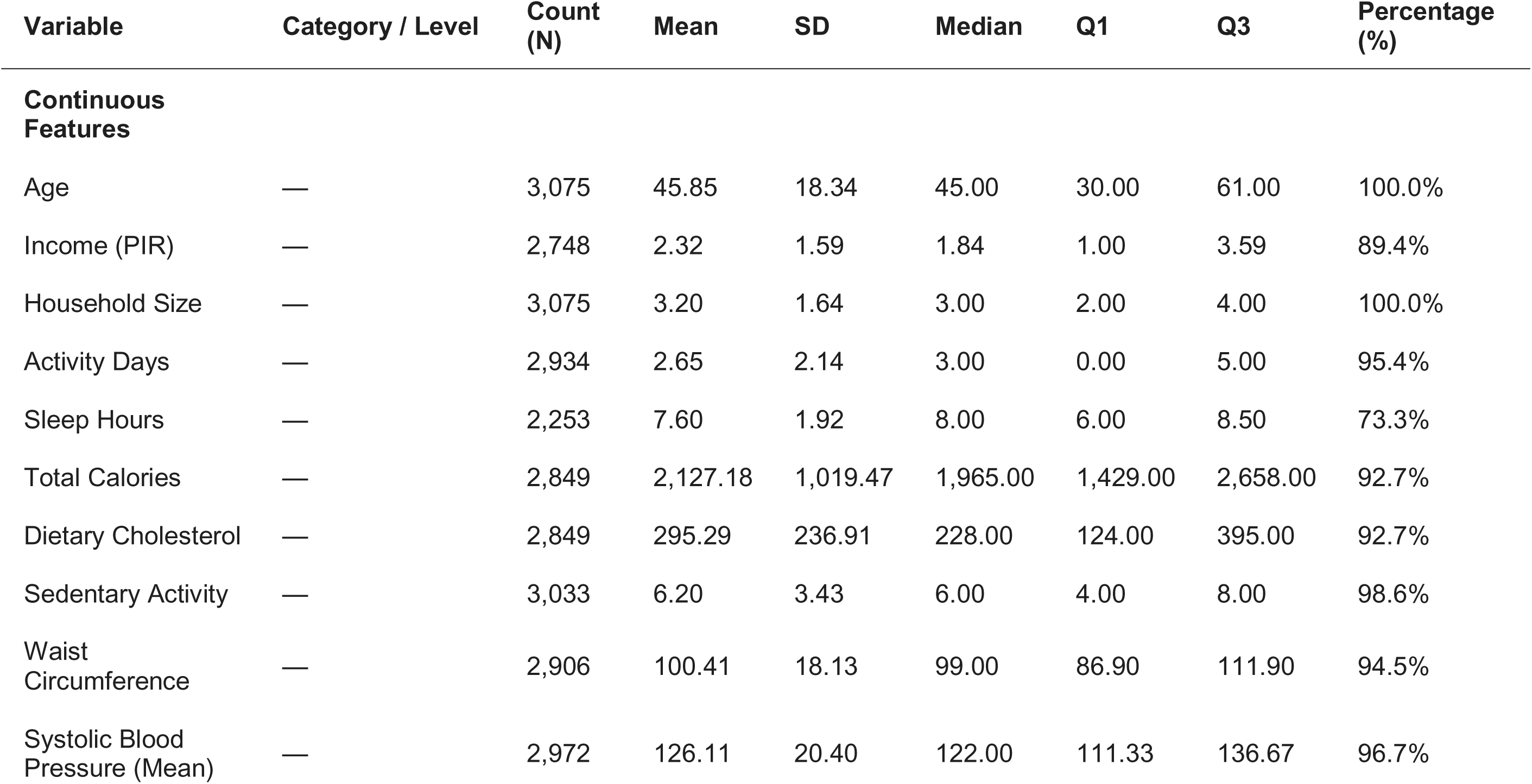
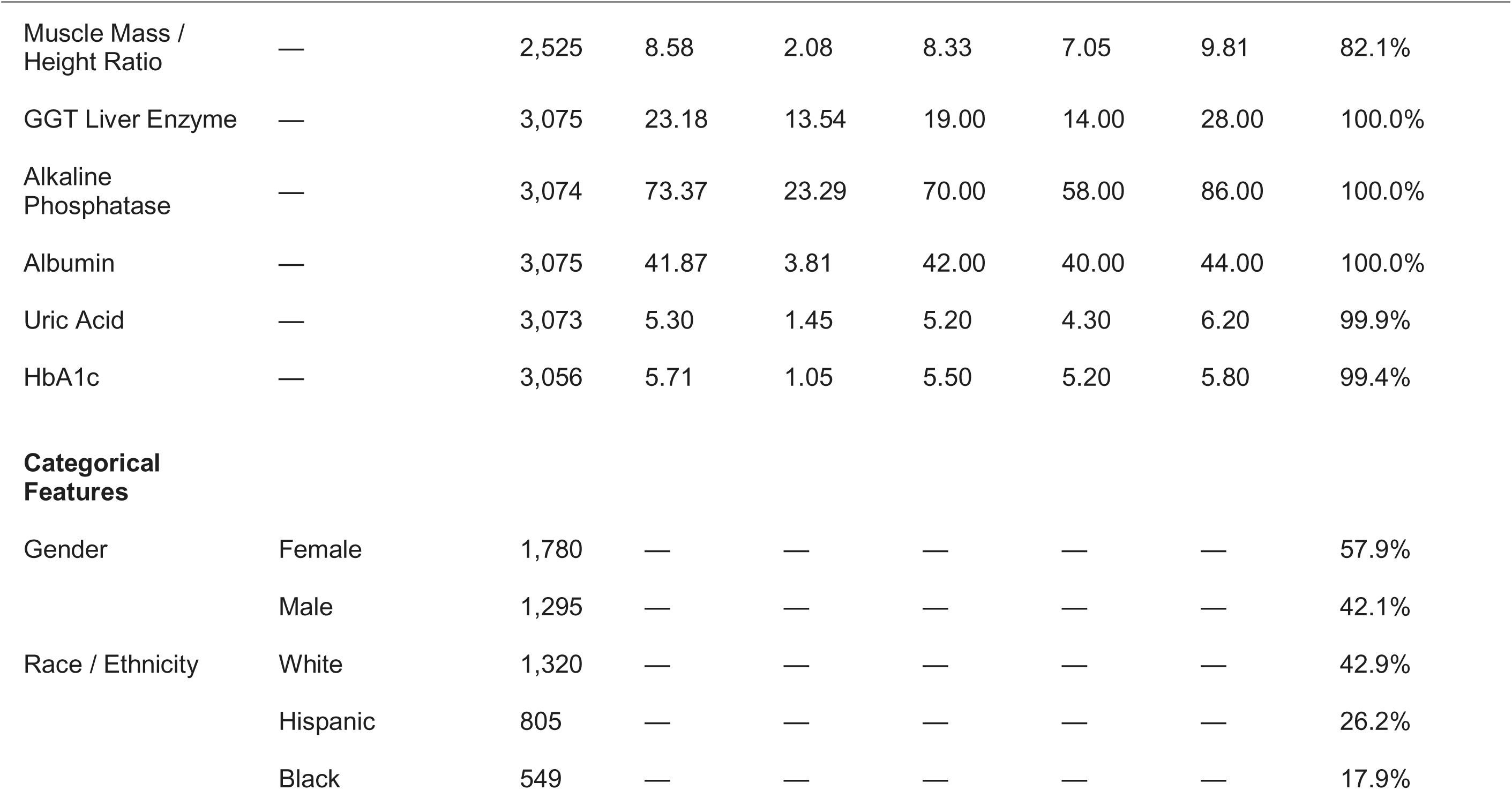
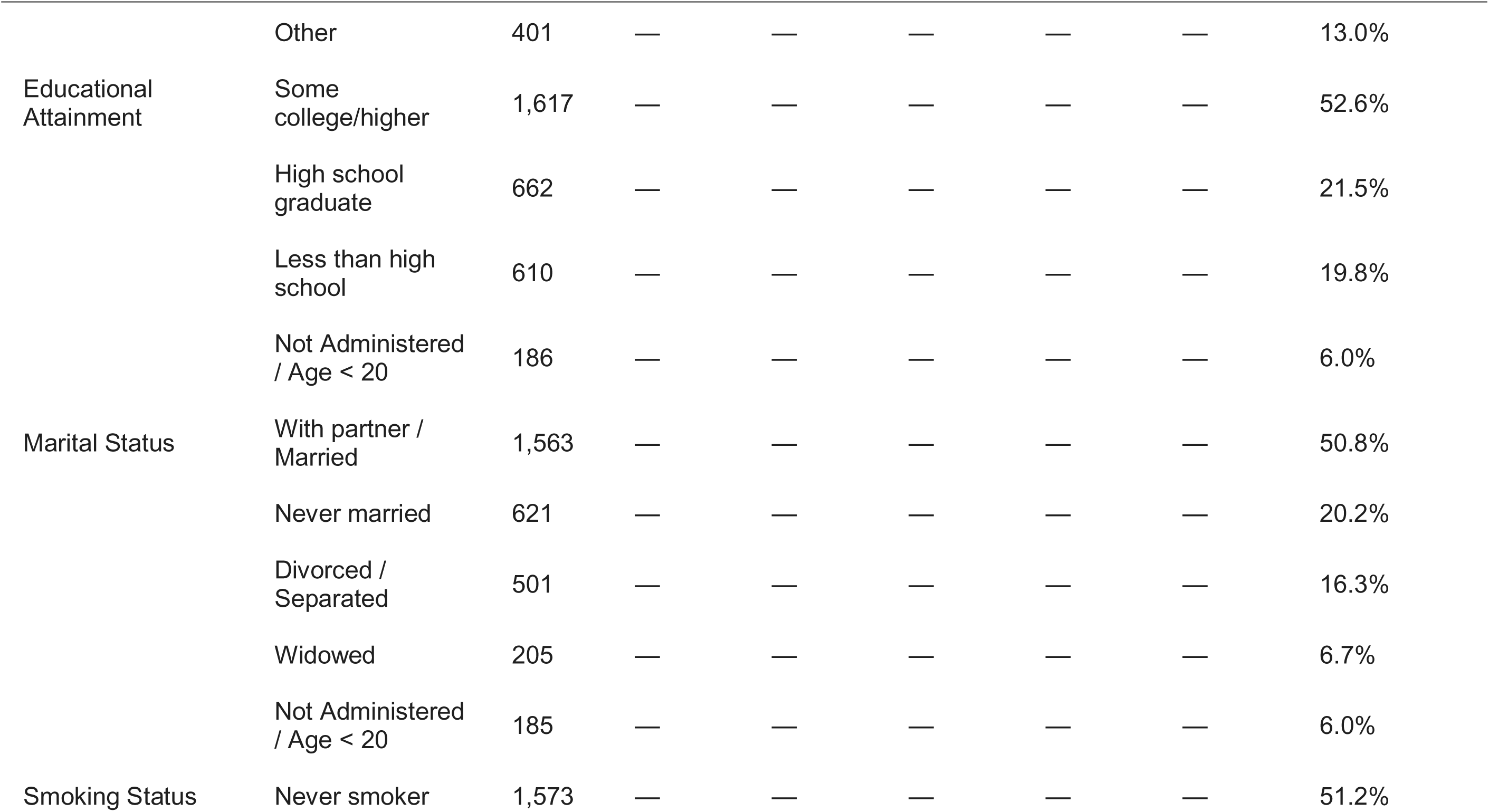
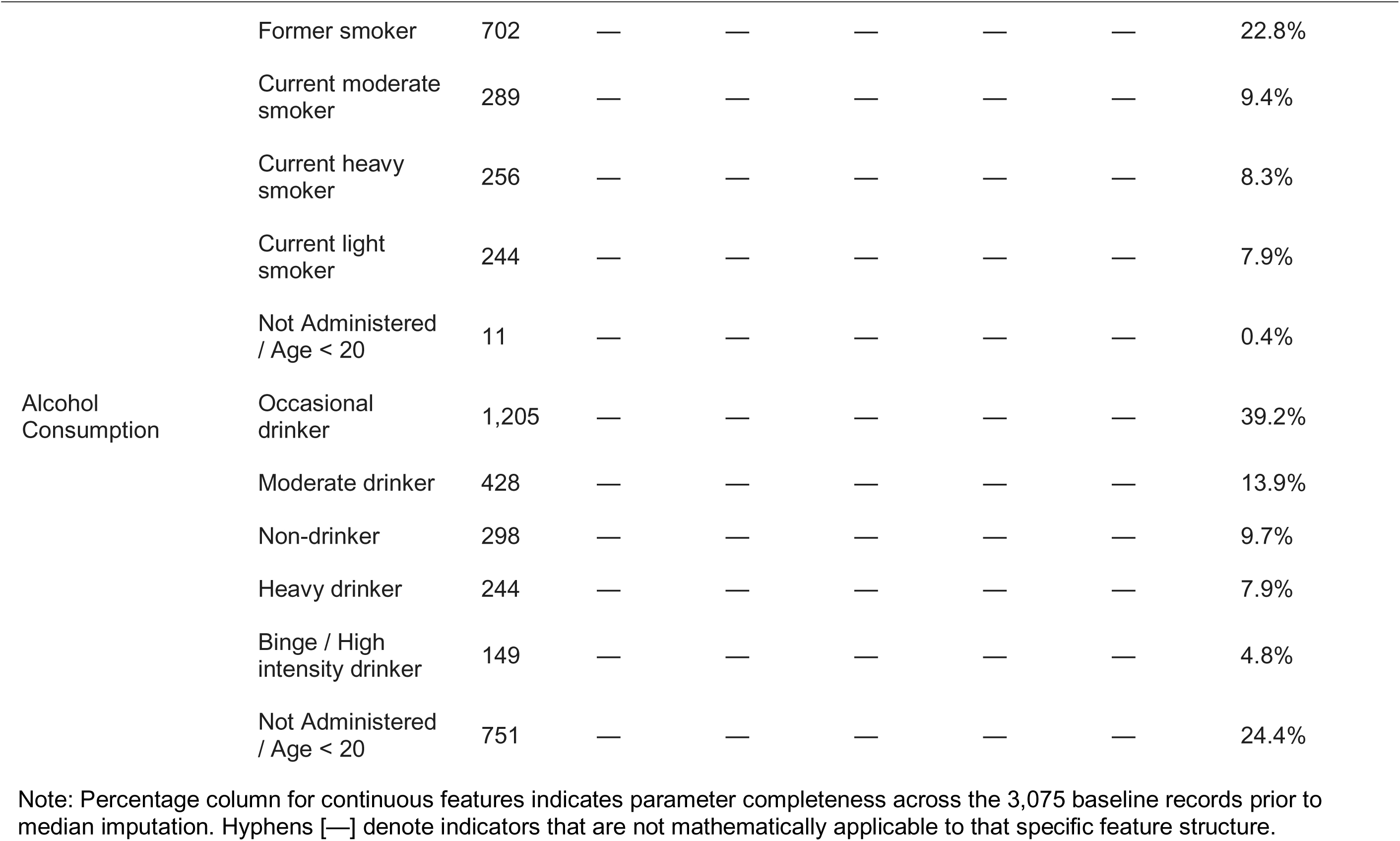
Baseline Demographic, Behavioural, and Physiochemical Characteristics of the Symptomatic Cohort (*N* = 3, 075)

### Sociodemographic and Social Infrastructure Profiles

The mean age of the symptomatic cohort was 45.85 ± 18.34 years. The sample skewed female (57.9%) relative to male (42.1%). In terms of racial and ethnic distribution, White participants comprised the largest segment (42.9%), followed by Hispanic (26.2%), Black (17.9%), and individuals identified as Other (13.0%).

Socioeconomic and structural vulnerabilities were prominent throughout the cohort:

- **Income Deprivation:** The mean Poverty Income Ratio (PIR) was 2.32 ± 1.59, with a median of 1.84. This indicates that over half of the symptomatic population lived below twice the federal poverty line, highlighting significant economic vulnerability.
- **Household Structure:** Participants lived in households averaging 3.20 ± 1.64 persons.
- **Administrative Gap:** Mirroring NHANES branching logic, 6.0% of the dataset (*n* = 186) fell within the late-adolescent 18-19 age bracket. Consequently, their Education and Marital Status records were coded as “Not Administered”. Among adult participants aged ≥ 20 years, 52.6% had completed some form of higher education, while 50.8% reported living with a spouse or partner.

### Behavioural and Lifestyle Characteristics

The behavioural baseline reflected substantial disruptions in sleep, physical activity, and substance use:

- **Sleep and Sedentary Patterns:** Participants reported a mean sleep duration of 7.60 ± 1.92 hours per day. However, they spent an average of 6.20 ± 3.43 hours per day in sedentary behavior.
- **Physical Immobility:** Physical activity levels were notably low; the median number of active days per week involving moderate-to-vigorous physical activity was 3.00, with a substantial portion of the population recording 0.00 active days (*Q*1 = 0.00).
- **Substance Use:** Active smoking was highly prevalent: 25.6% of the cohort were current smokers, divided across heavy (8.3%), moderate (9.4%), and light (7.9%) use patterns. Alcohol tracking revealed that 12.7% of the symptomatic population engaged in heavy or high-intensity binge drinking, while 24.4% missed the alcohol administrative screening due to age-related survey restrictions.

### Anthropometric and Physiochemical Strain

Physical health metrics indicated a population under chronic systemic and metabolic stress, validating the biopsychosocial overlap:

- **Visceral Obesity:** The mean waist circumference was 100.41 ± 18.13 cm (median: 99.00 cm). This places the average cohort member into the clinically high-risk category for metabolic syndrome.
- **Cardiovascular Strain:** The mean adjusted systolic blood pressure was elevated at 126.11 ± 20.40 mmHg, with the upper quartile reaching 136.67 mmHg, reflecting pervasive unmanaged or medication-masked hypertension.
- **Glycemic Control:** Long-term glycemic tracking via HbA1c revealed a mean of 5.71 ± 1.05%, indicating a baseline bordering on clinical pre-diabetes.
- **Skeletal Muscle Status:** The estimated skeletal muscle-to-height index (SMI) averaged 8.58 ± 2.08 kg/m^2^, establishing a clear physical baseline for downstream cluster comparisons.

### National Mental Health Prevalence and Treatment Gaps (Baseline Epidemiology)

To establish a nationwide baseline before applying advanced unsupervised cluster modelling, macro-epidemiological tracking was performed on the full population (*N* = 11,848). The cross-sectional analysis revealed a high symptom prevalence alongside treatment gaps spanning socioeconomic and demographic boundaries (Figure 1).

**Figure 1.**
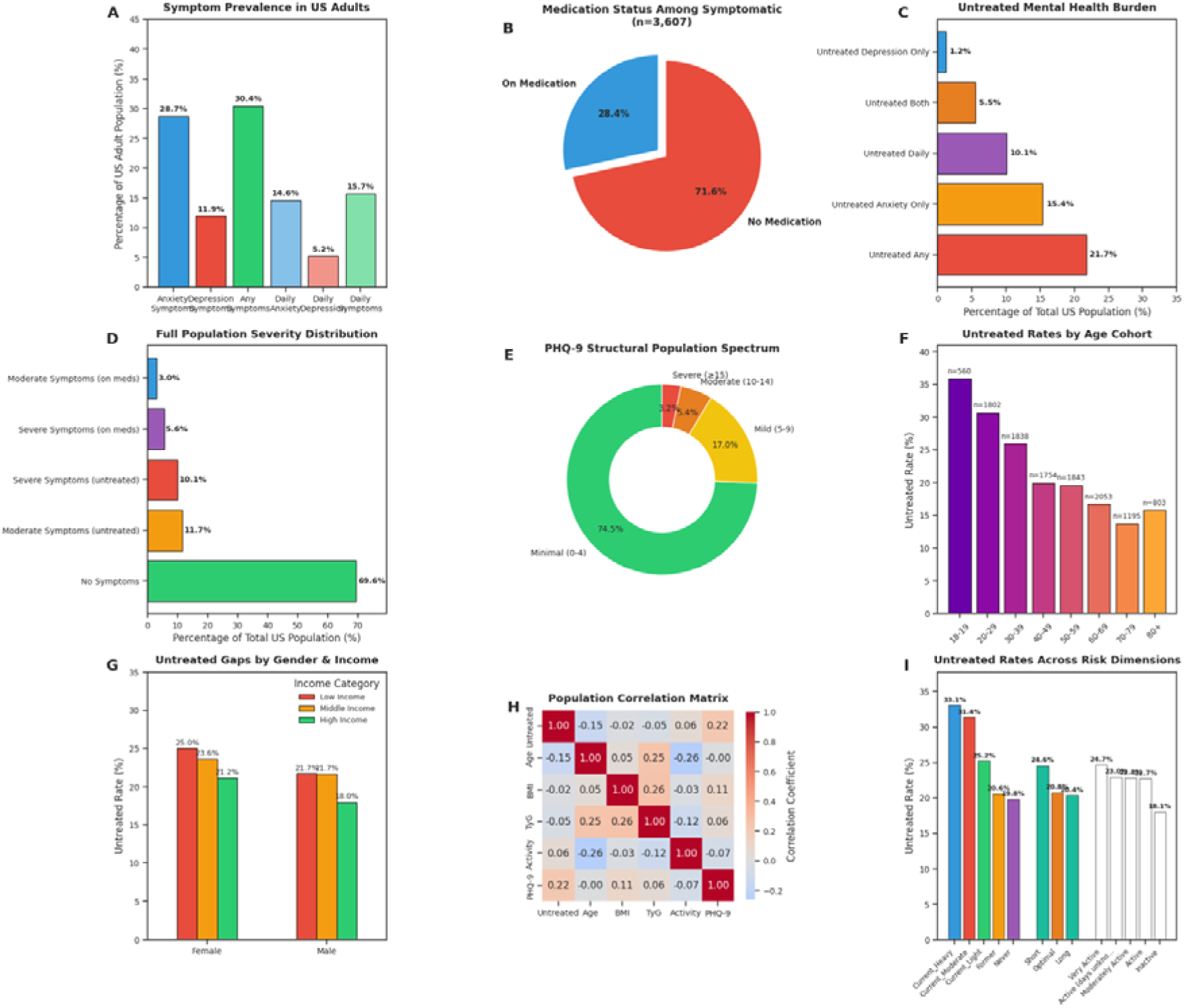
Macro-epidemiological landscape of mental health symptom prevalence, severity, and clinical treatment gaps among US adults. (A) Nationwide daily and weekly symptom prevalence; (B) Medication adherence vs. treatment deficit among the symptomatic sub-cohort ($n = 3,075); (C) Structural distribution of the untreated symptom burden; (D) Full population severity classification; (E) Population spectrum of clinical depression via PHQ-9 scoring; (F) Untreated population rates stratified across age cohorts; (G) Interactive disparities of untreated rates across gender and income categories; (H) Correlation matrix of primary continuous metrics; (I) Untreated rates mapped across behavioural risk and lifestyle dimensions.

### Broad Symptom Prevalence and Severity Distribution

The population baseline indicated a profound psychiatric burden across US adults. Nearly one-third of the population (30.4%) reported experiencing active symptoms of anxiety or depression on a daily or weekly basis.

- **Symptom Stratification:** Daily or weekly anxiety symptoms were highly prevalent, affecting 28.7% of the population, while depression symptoms affected 11.9%.
- **Daily Impairment:** Chronic, daily psychiatric distress was recorded in 15.7% of all respondents, driven primarily by daily anxiety (14.6%).
- **Clinical Severity and Validation:** When evaluated against the structured clinical PHQ-9 depression scale, 8.6% of the total population met the criteria for moderate-to-severe clinical depression (PHQ-9 score ≥ 10), demonstrating an 89.3% diagnostic concordance with the broad self-reported tracking metrics. Overall, 21.8% of the entire US adult population was found to be living with actively unmedicated moderate-to-severe psychiatric symptoms (11.7% moderate untreated; 10.1% severe untreated).

### Macro Treatment Gap

Among the individuals in the final analytical cohort (*N* = 3,075), an overwhelming majority were entirely disconnected from clinical pharmaceutical management. Of these symptomatic sufferers, 71.6% reported taking no medication for their distress, leaving a sparse 28.4% treatment rate across the modeled cohort.

When evaluated as a proportion of the entire US population, 21.7% of all adults represent an entirely untreated clinical burden. This untreated deficit was heavily characterized by isolated anxiety profiles (15.4%) and comorbid anxiety-depression presentations (5.5%).

### Demographic and Risk Factor Stratification

Stratifying the untreated population rate across demographic and lifestyle axes exposed critical disparities in healthcare access:

- **Vulnerability of Emerging Youth:** The untreated rate demonstrated a linear relationship with age. Late adolescents aged 18-19 exhibited the highest untreated rate at 36.0% (*n* = 560). The untreated rate was lower among middle-aged strata before tracking slightly higher in the elderly cohort (80+ years).
- **Socioeconomic Gaps:** Income and gender intersected to compound treatment disparities. Across both genders, individuals in low-income brackets experienced the highest untreated rates, peaking among low-income females at 25.0%.
- **Behavioural and Substance Use Risks:** The untreated burden was heavily concentrated among individuals with high-risk lifestyle habits. Current heavy tobacco users and current moderate smokers exhibited the highest overall untreated rates at 33.1% and 31.4%, respectively. Similarly, individuals with suboptimal sleep duration categories experienced elevated untreated rates (24.6%), underscoring a prominent overlap between behavioural risk profiles and a lack of clinical intervention.

## Latent Class Model Optimization and Selection

To establish the optimal number of sub-populations within the symptomatic cohort (*N* = 3,075), we evaluated the fit of the Gaussian Mixture Model using both the Akaike Information Criterion (AIC) and Bayesian Information Criterion (BIC) across a range of 2 to 6 components (Figure 2). Both metrics balance model likelihood against parameter complexity, with lower scores indicating superior fit without overparameterization.

**Figure 2.**
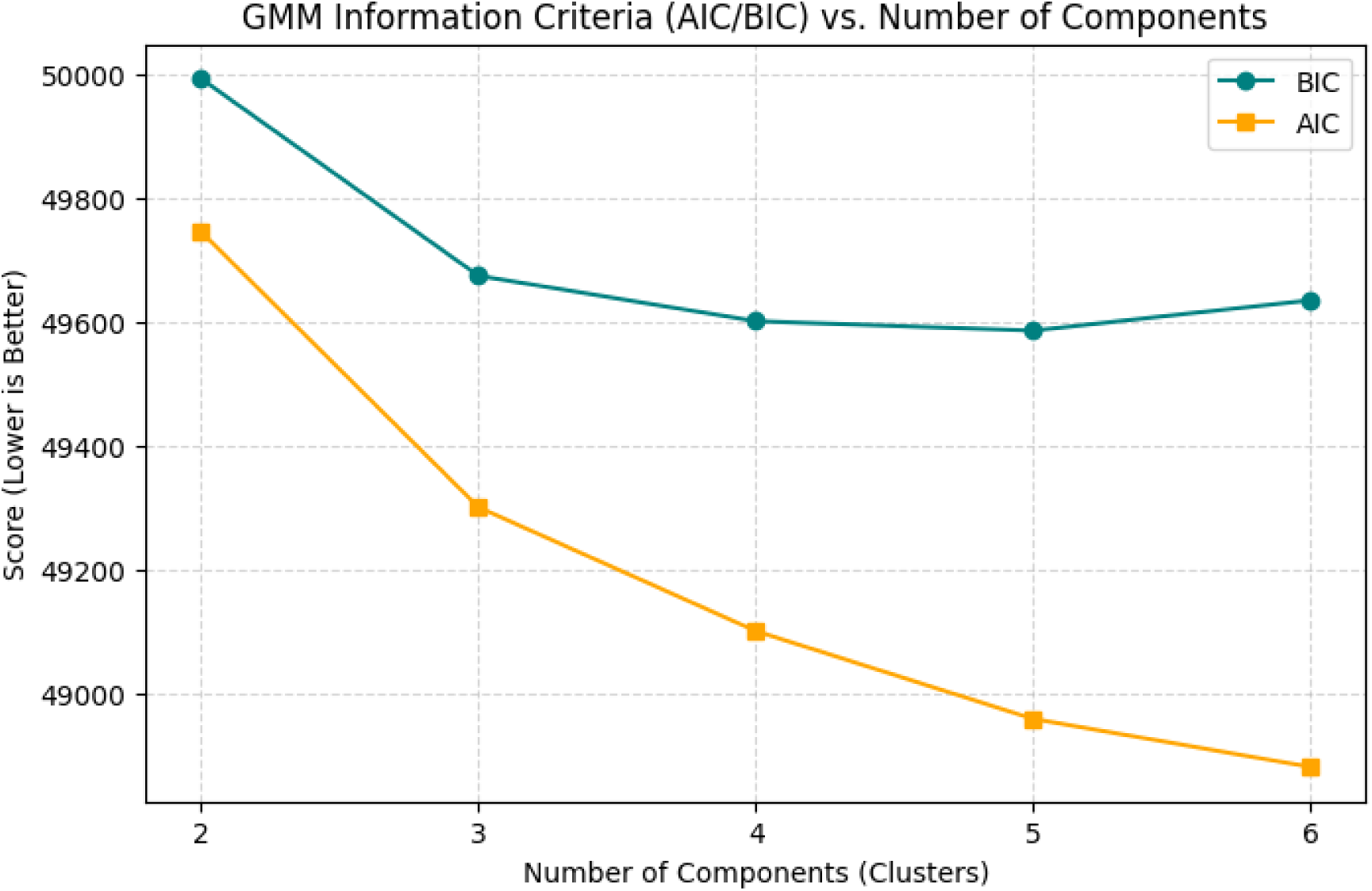
Gaussian Mixture Model (GMM) selection metrics across a 2-to-6 component range. The teal curve represents the Bayesian Information Criterion (BIC), which reaches a distinct structural inflection point (elbow) at 3 components before flattening and reversing at 6 components, indicating the onset of overfitting. The orange curve represents the Akaike Information Criterion (AIC).

### Information Criteria Analysis

The evaluation metrics demonstrated a distinct trajectory across the component range:

- **Two-Component Configuration:** The baseline structure yielded the highest error metrics, with a BIC of 49,992.82 and an AIC of 49,745.55.
- **Three-Component Configuration:** Moving from 2 to 3 clusters resulted in a sharp drop in both scores, with the BIC falling to 49,674.48 and the AIC dropping to 49,300.55. This substantial reduction signals a marked increase in explanatory variance.
- **Diminishing Returns (The Elbow):** Beyond 3 components, model fit improvement slowed considerably. The BIC curve flattened between 3 and 5 components (BIC = 49,601.23 at *k* = 4; BIC = 49,585.92 at *k* = 5).
- **Overfitting Boundary:** At 6 components, the BIC reversed direction and increased (BIC = 49,634.91), indicating the onset of overparameterization.

### Final Model Selection

While the more lenient AIC continued a gradual downward trajectory through 6 components (AIC = 48,881.02), we prioritized the more conservative BIC in accordance with standard statistical parsimony principles for large human samples to prevent cluster fragmentation. The distinct “elbow” at 3 components on the BIC curve represents the optimal balance of statistical power and parsimony. Consequently, a 3-cluster framework was selected as the definitive structure for downstream phenotypic profiling.

### Factor Analysis of Mixed Data (FAMD) Feature Loadings and Topology

To interpret the underlying latent structure guiding the clustering algorithm, feature loading scores for the first two principal components were analyzed. These orthogonal dimensions reduce the 22-dimensional feature matrix into a simplified coordinate space while preserving the primary variance of the mixed-type data. When projected in two dimensions, the GMM clusters exhibited distinct spatial segmentation along both axes (Figure 3).

**Figure 3.**
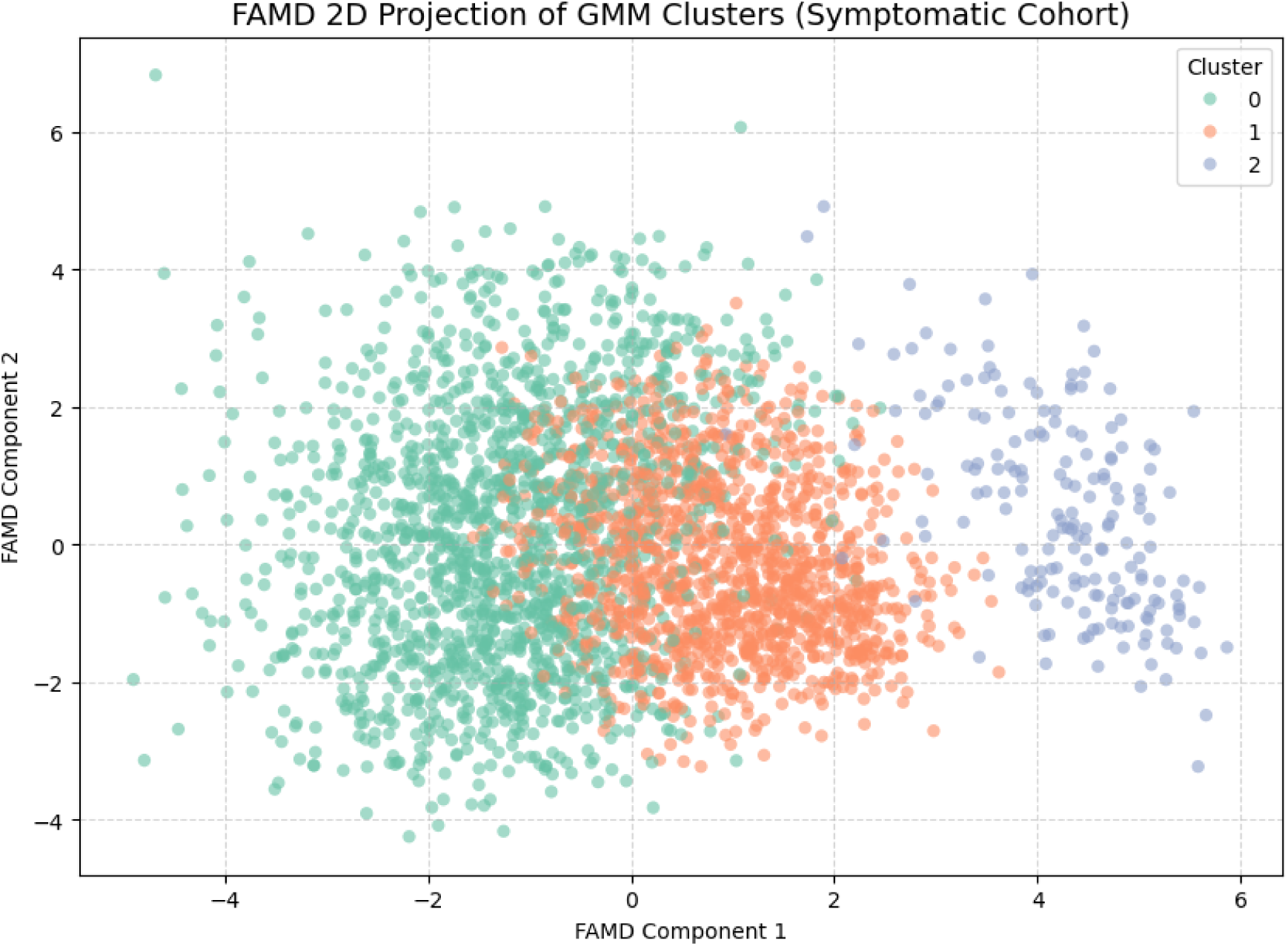
Two-dimensional projection of the symptomatic cohort across the first two FAMD principal components. Individual data points are mapped according to their latent coordinates and color-coded by their final Gaussian Mixture Model (GMM) cluster assignment. Cluster 0 represents the mature cardiometabolic cohort; Cluster 1 identifies the working-age professional segment; Cluster 2 isolates the highly vulnerable emerging youth pocket.

### Component 1: The Chronological and Cardiometabolic Axis

FAMD Component 1 served as a primary marker of chronological aging alongside its co-occurring biological and social transformations.

- **Negative Pole (Aging and Systemic Strain):** The negative end of this axis was heavily dominated by continuous physical health markers. Strong negative loadings were driven by Age (-0.751), Systolic Blood Pressure (-0.611), Waist Circumference (-0.546), and HbA1c (-0.483). This pole establishes a clear physiological gradient tracking advancing metabolic and cardiovascular wear-and-tear.
- **Positive Pole (Social Milestones and Early Life Stages):** Conversely, the positive pole was defined by institutional social variables, including Marital Status (0.473) and Education (0.386). Because the 18-19 age bracket lacks adult marital and educational markers due to NHANES survey logic, their records skewed sharply toward the far positive side of this axis.

### Component 2: The Physical Composition and Energy Throughput Axis

FAMD Component 2 captured independent variance related to body composition, sex-linked baseline physiology, and physical scale, running perpendicular to the chronological age axis.

- **Positive Pole (High Lean Mass and Intake):** The positive end of Component 2 was driven by structural physical traits, led by Muscle Mass / Height Ratio (0.769), Uric Acid (0.534), Total Calories (0.518), and Gender (0.447). This coordinate maps individuals with larger frames, higher lean skeletal indexes, and elevated caloric and metabolic throughput.
- **Negative Pole (Lower Metabolic Baseline):** The negative spectrum was comparatively flat, with minor contributions from behavioural recovery parameters such as Sleep Hours (-0.157).

### Latent Space Partitioning and Topographic Distribution

Visualizing the probabilistic GMM assignments within the 2D FAMD coordinate projection revealed clear structural separation of the symptomatic population (Figure 3):

- **Cluster 0 (Green):** Concentrated almost entirely on the negative end of Component 1 (spreading from approximately -5 to 0), this cluster mapped tightly along the chronic biological stress and aging pole.
- **Cluster 1 (Orange):** Positioned centrally along Component 1 but stretching vertically across the mid-to-lower sections of Component 2, this large sub-population represented a mid-life cohort defined by high workforce lifestyle indicators and healthy skeletal muscle mass profiles.
- **Cluster 2 (Blue):** Strikingly isolated on the extreme positive end of Component 1 (spanning from +2 to +6), this group formed a highly condensed, structurally distinct cluster. Cluster 2 emerged as a distinct, tightly condensed topology due to the unique variance profile of the 18-19 age bracket within the FAMD coordinate space.

## Socio-Demographic and Behavioural Phenotypes of the Latent Clusters

To characterize the demographic and behavioural profiles defining each sub-population, cross-tabulated distributions were evaluated across the three optimized GMM clusters (Figure 4).

**Figure 4.**
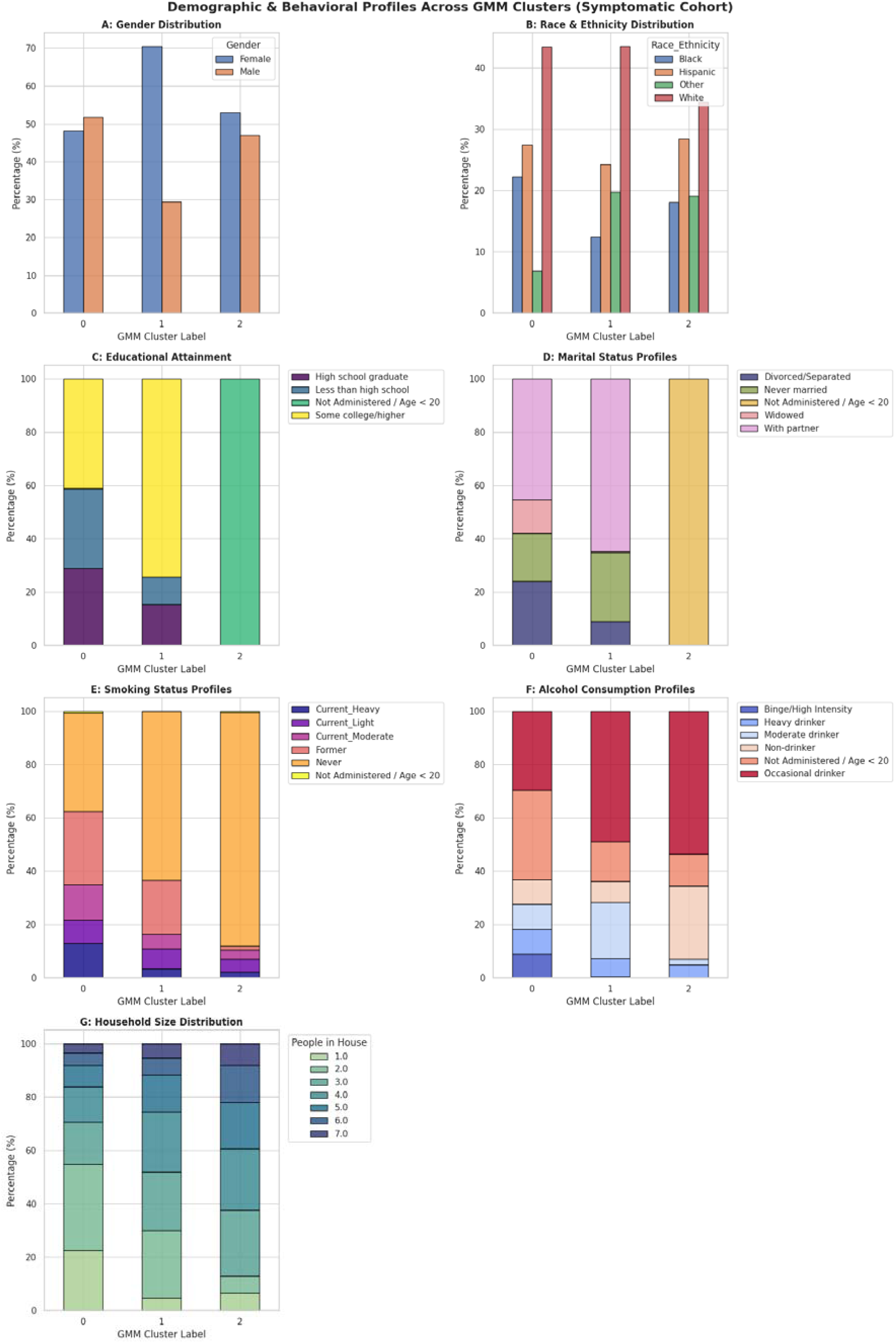
Demographic, institutional, and behavioural risk profiles stratified across the three optimized latent GMM clusters (. (A) Gender distribution showing the high female concentration in Cluster 1; (B) Racial and ethnic composition across segments; (C) Educational attainment highlighting the structural “Under 20” administrative block in Cluster 2; (D) Marital status distribution tracking social isolation in Cluster 0; (E) Smoking status profiles highlighting severe tobacco exposure in Cluster 0; (F) Alcohol consumption typography; (G) Household size distributions tracking domestic density.

The analysis showed that the machine learning pipeline separated the broad symptomatic population into highly distinct life-stage and socio-behavioural risk cohorts.

### Cluster 0: Mature, High-Burden Cohort

Cluster 0 represented a socio-economically vulnerable, mature adult segment experiencing high chronic behavioural and systemic risk exposures.

- **Demographics and Socioeconomic Standing:** This cohort displayed a balanced gender distribution (48.2% female vs. 51.8% male) and a strong concentration of White (43.4%) and Hispanic (27.4%) participants. Educational attainment was notably compressed compared to the other adult cluster, with 58.7% of the group possessing a high school education or less (28.9% high school graduate; 29.8% less than high school).
- **Social Architecture:** Social isolation markers were uniquely pronounced in this cluster. While 45.4% were married or living with a partner, this cluster held the highest proportion of divorced or separated individuals (24.1%) and widowed participants (12.6%). Correspondingly, over half of the cohort resided in single- or two-person households (22.6% single; 32.3% dual occupancy).
- **Behavioural Risk Profiles:** This group carried a notably heavy substance use burden. Active tobacco use peaked sharply in Cluster 0, with a combined 34.9% current smoking rate (13.0% heavy; 13.2% moderate; 8.7% light). High-intensity alcohol consumption was also heavily represented, with 18.2% categorized as active heavy or binge/high-intensity drinkers.

### Cluster 1: High-Functioning, Workforce Professional Cohort

Cluster 1 identified a large, highly educated, middle-aged segment characterized by stable social frameworks but widespread behavioural masking of symptoms.

- **Demographics and Diversity:** This group was heavily dominated by female participants (70.5%) and exhibited a diverse racial profile (43.5% White, 24.3% Hispanic, 12.4% Black, and 19.8% Other). It represented an economically and socially integrated segment of the workforce.
- **Socioeconomic and Social Capital:** Educational and social capital were exceptionally high. Nearly three-quarters of this cohort (74.2%) had attained some form of higher education. Structurally, 64.7% maintained stable domestic partnerships, and family units were substantially larger, with 48.0% residing in households with 4 to 7 people.
- **Lifestyle and Coping Patterns:** In contrast to Cluster 0, this cohort demonstrated low smoking engagement, with 63.3% reporting as never-smokers. Alcohol patterns showed a shift toward social or occasional drinking, with 49.0% classified as occasional drinkers and only 7.1% meeting the criteria for regular heavy or binge consumption.

### Cluster 2: Disconnected Emerging Youth Cohort

Cluster 2 isolated a compact, structurally distinct, and highly vulnerable late-adolescent sub-population (18-19 years of age) facing severe systemic barriers to care.

- **Administrative Trap:** Due to NHANES survey branching pathways, 100.0% of this cohort’s data for formal adult educational attainment and marital status profiles was skipped and recorded as “Not Administered”. This group represented a cohort transitioning into early adulthood without traditional domestic or institutional frameworks.
- **Demographics and Household Structures:** The gender split was relatively even (53.0% female vs. 47.0% male), with prominent representation of Black (18.0%) and Hispanic (28.4%) youth. They lived in densely populated, multi-generational domestic spaces; only 6.6% lived alone, while 62.3% resided in large households containing 4 to 7 people.
- **Behavioural Gaps:** Substance use markers indicated early-stage exposures, with an 87.4% never-smoker rate and a high concentration of occasional drinkers (53.6%). Notably, 27.3% missed the alcohol usage screening entirely due to age-related survey restrictions, confirming the unique administrative footprint that allowed the GMM to isolate this vulnerable pocket.

## Continuous Feature Variation and Statistical Significance across Latent Classes

To evaluate the relative contribution of each biological and social metric to cluster differentiation, an Analysis of Variance (ANOVA) was performed. Each continuous feature was ranked according to its F-statistic (a metric evaluating the ratio of variance between the clusters to the variance within them) to measure the strength of feature differentiation across the cohorts (Figure 5). A higher F-statistic indicates that a variable varies significantly between distinct clusters while remaining tightly grouped within each individual cluster. The analysis confirmed a strong biological gradient, with physical health degradation tracking closely with distinct life-stage cohorts.

**Figure 5.**
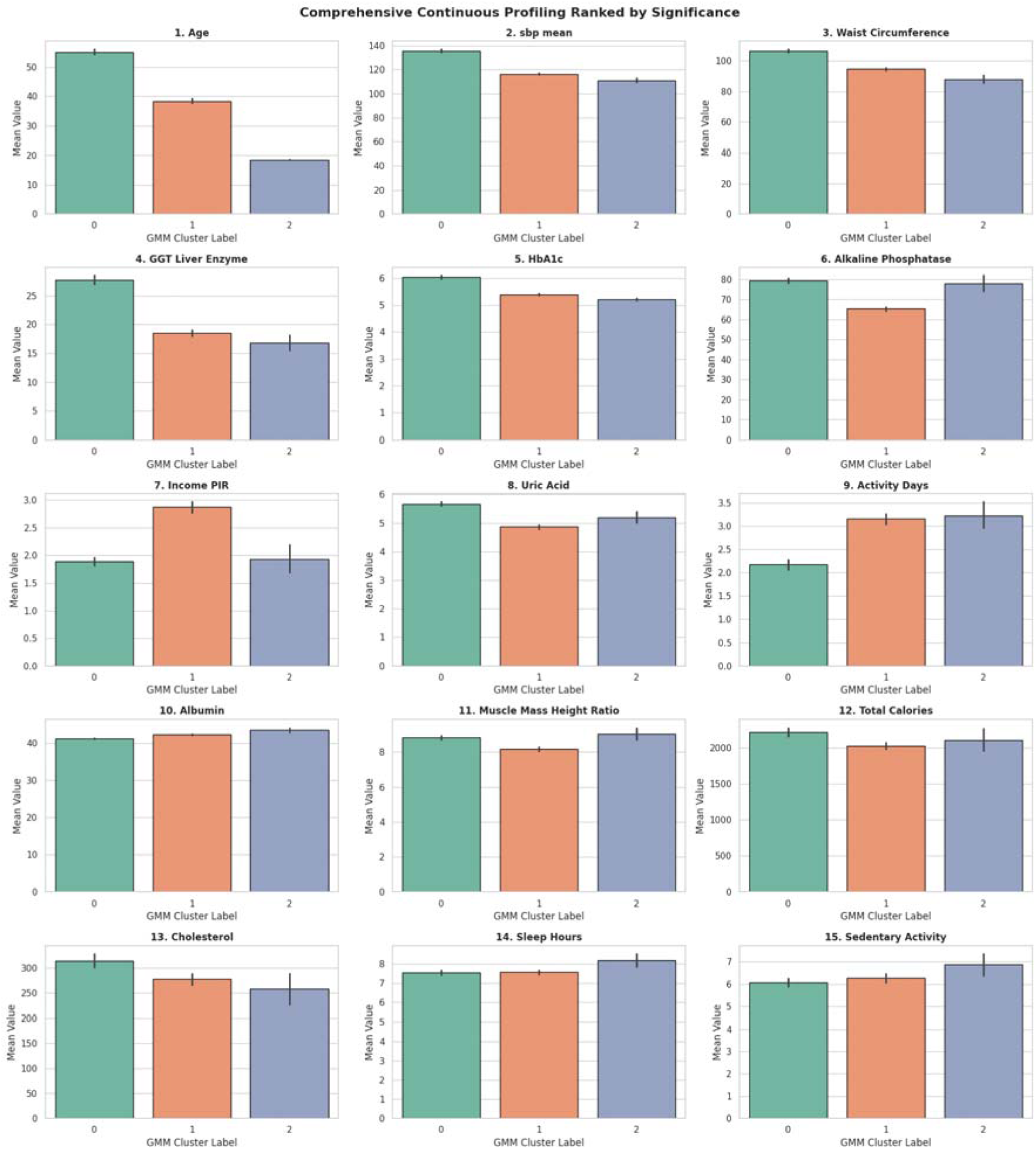
Comprehensive continuous feature profiling across latent GMM clusters, ranked in descending order of statistical significance (ANOVA F-statistic). Bars represent mean values for each sub-population, with vertical error bars indicating confidence intervals. Panels 1–6 identify the primary biological and clinical biomarkers of aging and metabolic decay; panels 7–11 map out intersecting socioeconomic and physical vitality markers; panels 12–15 highlight low-significance behavioural habits that remain largely consistent across the entire symptomatic cohort.

### 1. High-Significance Diagnostic Vectors (*F* > 150)

The primary drivers of cluster separation were chronological age and downstream clinical biomarkers of cardiovascular, liver, and metabolic wear-and-tear:

- **Age (***F* = 776.5): This parameter served as the strongest divider in the dataset. The mean ages across the groups highlighted a clear lifespan progression: Cluster 0 captured a late-adulthood cohort (55.03 years), Cluster 1 represented middle-aged adults (38.38 years), and Cluster 2 isolated an emerging youth pocket (18.49 years).
- **Cardiovascular Strain (***F* = 488.6): Adjusted systolic blood pressure (sbp_mean) dropped in a step-wise pattern across the age spectrum. Cluster 0 exhibited severe, chronic hypertensive strain (135.82 mmHg), Cluster 1 matched early pre-hypertensive baselines (116.27 mmHg), and Cluster 2 tracked along a youthful baseline (111.15 mmHg).
- **Visceral Adiposity (***F* = 217.6): Mean waist circumference varied substantially between the groups. Cluster 0 presented severe clinical central obesity (106.64 cm), which dropped to a moderate threshold in Cluster 1 (94.69 cm) and reached its lowest levels in Cluster 2 (88.16 cm).
- **Hepatic and Glycemic Deterioration:** Both GGT_Liver_Enzyme (*F* = 211.6) and long-term glycemic control HbA1c (*F* = 167.5) reached their worst clinical metrics in Cluster 0 (GGT = 27.70 U/L; HbA1c = 6.03%), establishing a high pre-diabetic risk threshold. These values settled substantially in Cluster 1 (5.40%) and Cluster 2 (5.21%).

### 2. Moderate-Significance Socioeconomic and Systemic Drivers (*F* = 30 to 150)

Socioeconomic capital, metabolic byproducts, and muscle composition formed the secondary tier of cluster differentiation:

- **Socioeconomic Resource Gaps (***F* = 140.2): Financial capital was highly concentrated in the middle-aged workforce segment (Cluster 1), which recorded a peak mean Poverty Income Ratio of 2.87. Conversely, Cluster 0 (1.89) and Cluster 2 (1.93) were locked into near-identical low-income profiles, demonstrating that different life stages can share similar economic limitations.
- **Physical Activity Baselines (***F* = 83.1): Middle-aged professionals (Cluster 1) and emerging youth (Cluster 2) maintained similar physical activity habits (3.15 and 3.22 active days per week, respectively), while the mature Cluster 0 cohort was significantly more immobile (2.17 days).
- **Skeletal Muscle Index (***F* = 34.3): Despite being decades younger, the emerging youth of Cluster 2 maintained a high lean physical frame with a Muscle_Mass_Height_Ratio of 9.03 kg/m^2^, comparable to the mature Cluster 0 cohort (8.83 kg/m^2^) and exceeding the female-dominated Cluster 1 professional group (8.16 kg/m^2^).

### 3. Low-Significance Lifestyle Invariance (*F* < 15)

Parameters including total daily caloric intake (*F* = 11.4), dietary cholesterol (*F* = 10.4), sleep duration (*F* = 6.9), and sedentary hours (*F* = 4.9) showed minimal variation across the three groups. While Cluster 2 youth recorded slightly higher sleep duration (8.19 hours), the entire symptomatic population remained uniformly sedentary, averaging 6.07 to 6.87 hours of sitting time per day regardless of cluster membership.

## Clinical Severity and Treatment Gaps Across Clusters

To examine the relationship between life stage and medication non-adherence, clinical psychiatric severity and treatment status were cross-tabulated across the three GMM clusters (Figure 6). This analysis revealed the structural nature of the untreated burden, demonstrating that the treatment gap varied substantially across different stages of life.

**Figure 6.**
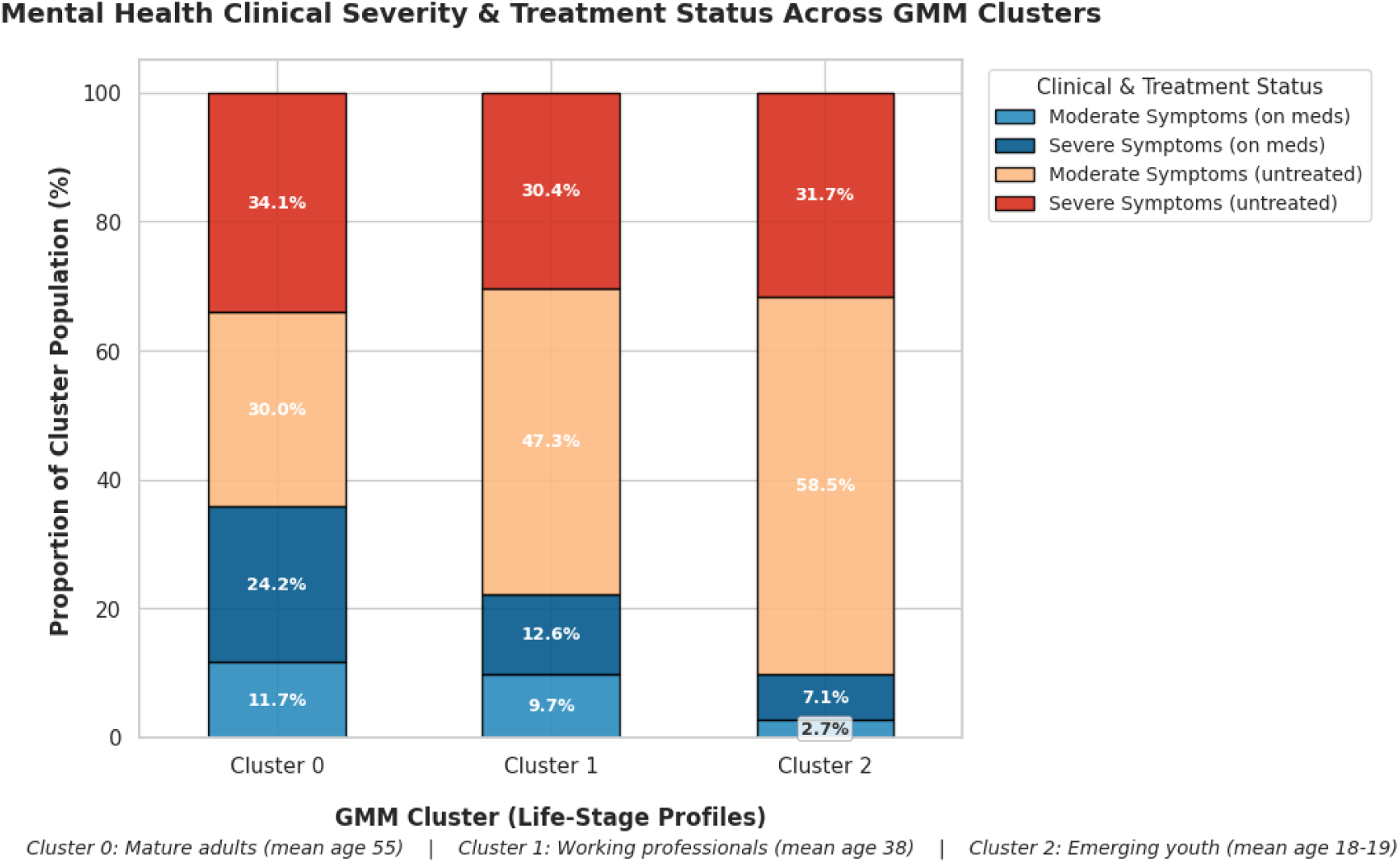
Distribution of mental health clinical severity and pharmaceutical treatment status stratified across the three latent life-stage GMM clusters (. Stacked bars illustrate the proportion of each sub-population falling into four distinct clinical categories. Blue shading indicates active pharmaceutical treatment compliance; orange and red segments identify the untreated population burden. The graph illustrates a step-wise increase in the total untreated deficit as the cohort shifts from mature adulthood (Cluster 0, untreated) to middle-aged workforce professionals (Cluster 1, untreated) and reaches its peak among transitioning emerging youth (Cluster 2, untreated).

### Cluster 0: Mature Adults (Mean Age ∼55)

This cohort exhibited the highest overall level of severe psychiatric symptoms alongside the greatest rate of clinical pharmaceutical management. The group had the highest treatment rate in the study at 35.9% (11.7% moderate on medication; 24.2% severe on medication). Despite this, a substantial residual burden remains: 64.1% were entirely untreated, with 34.1% suffering from severe, unmedicated distress. This pattern suggests a clinical population that remains unreached by mental health services despite regular contact with primary care for chronic cardiometabolic conditions.

### Cluster 1: Working Professionals (Mean Age ∼38)

This cohort represented a substantial workforce segment managing psychiatric symptoms largely in clinical isolation. Moderate psychiatric symptoms affected 57.0% of the group (47.3% untreated; 9.7% on medication). The overall treatment deficit was high, leaving 77.7% of this cohort entirely unmedicated. The largest single block consisted of individuals experiencing moderate untreated symptoms (47.3%), a pattern consistent with high-functioning behavioural masking where individuals maintain professional and family roles while remaining disconnected from psychological or pharmaceutical support.

### Cluster 2: Emerging Youth (Aged 18-19)

This cohort isolated a highly vulnerable pocket experiencing near-total non-treatment, with a critical 90.2% overall untreated rate and only 9.8% connected to medical infrastructure. The vast majority managed active mental health issues without formal support: 58.5% experienced untreated moderate symptoms and 31.7% faced untreated severe symptoms. This stark treatment gap suggests that late adolescents transitioning away from paediatric or school-based support systems face unique structural barriers, potentially lacking the independent economic means, social structures, and healthcare navigation experience required to access adult mental health services.

## Discussion

This study demonstrates that the mental health treatment gap is a complex, multi-dimensional challenge that extends across geographic and cultural contexts. By applying a Factor Analysis of Mixed Data (FAMD) and Gaussian Mixture Modelling (GMM) framework to a symptomatic US cohort (*N* = 3,075), we isolated three distinct life-stage sub-populations facing unique socio-behavioural and biological profiles.

When compared to recent international data—notably the Singaporean study by Teo et al. (2026), which found that 77% of adults with mental health symptoms had not sought formal care—our findings reveal noteworthy parallels across populations.

Despite differences in healthcare infrastructure and cultural context between Singapore and the United States, the latent phenotypes associated with untreated psychiatric distress show similar structural patterns. By moving beyond broad population averages, our unsupervised machine learning approach provides a more granular understanding of the clinical, socioeconomic, and lifestyle profiles of untreated groups, offering a foundation for targeted public health strategies.

### Adolescent Transition and Cross-Cultural Vulnerability

A primary finding of this study is the isolation of Cluster 2 (Emerging Youth, mean age 18.49), a highly condensed and vulnerable segment experiencing substantial disconnection from psychiatric care (90.2% untreated). This finding aligns with demographic trends reported by Teo et al. (2026) in Singapore, where younger age cohorts similarly exhibited a higher symptom burden and lower service utilization.

Our data highlight a potential structural mechanism underlying this shared vulnerability: this cohort falls into a developmental and administrative gap. Every member of this cluster is aged 18-19, meaning that core sociological parameters such as educational attainment and marital status were omitted due to survey logic. In both Western and Asian contexts, this administrative gap reflects a critical life transition where young adults age out of paediatric or school-based support frameworks but may lack the financial independence (mean PIR = 1.93), social stability, and healthcare navigation experience required to access adult medical systems. With nearly a third of this group experiencing severe untreated symptoms (31.7%), this population may benefit from targeted early intervention.

### Symptom Masking and Barriers in the Working-Age Cohort

In contrast, Cluster 1 (Working Professionals, mean age 38.38) presents a different pattern of barriers to care. This female-dominated (70.5%), highly educated (74.2%), and financially stable (mean PIR = 2.87) cohort reported a 77.7% untreated rate, driven largely by untreated moderate symptoms (47.3%).

This pattern is consistent with the behavioural insights from Teo et al. (2026), which found that white-collar professionals and highly educated adults expressed interest in informal support channels over traditional clinical options. Cluster 1’s profile—stable relationships (64.7% married/partnered), active careers, and good physical health (HbA1c 5.40%, waist circumference 94.69 cm)—suggests behavioural masking: maintaining external functioning while managing symptoms privately. For this high-functioning cohort, barriers to care may include time constraints and concerns about professional stigma, though these factors were not directly measured in this study.

### Somatic Diversion and the Isolation of Primary Care

Finally, Cluster 0 (Mature Adults, mean age 55.03) was characterized by metabolic and cardiovascular strain, including central obesity (mean waist circumference 106.64 cm), elevated systolic blood pressure (135.82 mmHg), pre-diabetic glycemic profiles (HbA1c 6.03%), and heavy tobacco use (34.9%). While this group had the highest treatment rate in the study (35.9%), 64.1% remained unmedicated, with 34.1% experiencing severe untreated symptoms.

This profile raises the possibility of somatic diversion—whereby individuals in this life stage interact with healthcare systems for chronic disease monitoring while underlying psychiatric symptoms remain unaddressed. This pattern is consistent with the hypothesis that primary care clinicians may focus on managing physical biomarkers without systematically screening for comorbid mental health conditions, though this was not directly assessed in our study.

### Methodological Comparison and Analytical Strengths

Several methodological distinctions between this study and the Singaporean baseline by Teo et al. (2026) highlight the analytical contributions of our framework:

- **Sample Scale:** While the Teo et al. study provided valuable community insights from 350 participants, our larger, nationally representative cohort (*N* = 3,075 symptomatic individuals from an 11,848 baseline) provided the statistical power necessary for complex unsupervised clustering without model fragmentation.
- **Objective Biomarkers:** Unlike the Teo et al. framework and much of the psychiatric survey literature, our NHANES-derived dataset integrated objective physical examinations and biochemical laboratory data (including adjusted systolic blood pressure, visceral adiposity, HbA1c, and hepatic enzymes). Integrating objective biomarkers enabled post-hoc ANOVA testing to demonstrate a significant somatic gradient across the resulting latent classes (F-statistics up to 488.6), complementing subjective distress measures with objective health indicators.
- **Granular Life-Stage Tracking:** While the Singaporean study examined the adult population as a single block, our FAMD and GMM pipeline isolated distinct, narrow age bands—most notably Cluster 2, which captured late adolescents aged 18-19. This granularity highlighted a developmental and administrative transition that broader population averages may obscure.

### Cross-National Strategic Implications

The structural alignment between our US latent profiles and the Singaporean baseline data suggests that a uniform, clinical-first public health strategy may be insufficient to bridge the mental health treatment gap. Intervention frameworks could be tailored to the unique socio-behavioural traits of each life stage, utilizing both formal medical networks and informal community channels:

- **For Emerging Youth (Cluster 2):** Public health systems might consider developing confidential, mobile-first peer support networks that bypass traditional clinical settings. Late adolescents often show a preference for digital privacy and peer-led interaction, and such platforms could provide an accessible community bridge.
- **For Working Professionals (Cluster 1):** Addressing the treatment gap among this cohort could involve lowering logistical and reputational barriers to care. Employers and wellness programs might consider embedding virtual, anonymous peer-led support channels into corporate frameworks, mirroring the interest in informal networks observed by Teo et al. (2026).
- **For Mature Adults (Cluster 0):** Closing the gap for aging populations may require shifting away from siloed specialist care. Primary care and metabolic clinics could systematically co-locate mental health screening and behavioural counseling into routine chronic disease pathways, treating metabolic and psychiatric health as interconnected concerns.

### Limitations

Several limitations should be considered when interpreting these findings. First, the cross-sectional design of NHANES precludes causal inferences about the developmental trajectories suggested by the cluster structure. Second, the primary outcome was defined as pharmacologically untreated, excluding non-pharmacological interventions such as psychotherapy or peer support. Third, individuals with severe liver pathology (GGT ≥ 80 U/L) were excluded, which may have removed the most metabolically compromised individuals from the mature adult cluster. Fourth, missing data were present for several variables, most notably sleep (73.3% complete) and alcohol consumption (75.6% complete). Fifth, barriers to care—such as stigma, time constraints, and healthcare literacy—were inferred from demographic patterns rather than directly measured. Finally, while comparisons to international data are valuable, the cross-national inferences are based on a single comparison study and would benefit from replication across diverse settings.

## Conclusion

This study demonstrates the utility of unsupervised machine learning for uncovering hidden patterns within the mental health treatment gap. By applying an FAMD and Gaussian Mixture Modelling pipeline to a large, biomarker-enriched dataset, we showed that the unmedicated population is not a single, uniform group. Instead, it comprises distinct, life-stage-specific sub-populations: vulnerable transitioning youth affected by administrative gaps, high-functioning working professionals managing their symptoms without formal care, and mature adults whose severe psychiatric distress may remain unaddressed during routine physical healthcare.

These profiles help illustrate why traditional, one-size-fits-all healthcare strategies may fail to close the treatment gap. When compared to smaller, survey-based international data—such as the Singaporean study by Teo et al. (2026)—our findings reveal noteworthy parallels in how untreated mental illness manifests across the two populations examined. Ultimately, closing the mental health treatment gap may require shifting beyond broad population averages toward more targeted, community-driven interventions—including digital peer support networks for youth and integrated primary care for older adults—that address the unique clinical and social realities of each life stage.

## Declarations

### Ethical Approval / IRB

This study involved secondary analysis of publicly available, fully de-identified data from the Centers for Disease Control and Prevention (CDC) National Health and Nutrition Examination Survey (NHANES) cycles 2015-2018. As such, institutional review board approval was not required.

### Use of Artificial Intelligence

During the preparation of this manuscript, the author used generative AI tools for the purposes of code development and debugging, report organization, language refinement, and academic drafting assistance. All directing of analytical modelling and data interpretation, and final manuscript verification were performed exclusively by the human author, who accepts full accountability for the integrity of the work.

### Data Availability Statement

All data used in this study are publicly available from the CDC NHANES repository at https://www.cdc.gov/nchs/nhanes/.

